# Estimating the total morbidity burden of COVID-19

**DOI:** 10.1101/2021.04.20.21255818

**Authors:** Maia P. Smith

**Author notes:** (corresponding author), or 473 – 459 – 2693.

## Abstract

**Background:** Calculations of disease burden of COVID-19 are used to allocate scarce resources and historically have focused on mortality, with little attention to morbidity such as postviral ‘post-COVID,’ similar to chronic fatigue syndrome (CFS), which strikes 4 and 16% of male and female survivors. This paper quantifies post-COVID disability burden and combines it with case fatality to estimate total morbidity per COVID-19 case.

**Methods:** Healthy life years lost per COVID-19 case were computed as the sum of (incidence*disability weight*remaining lifespan) for death and post-COVID (modeled as CFS) by sex and 10-year age category. In addition to death, the main model considered lifelong mild, moderate or severe CFS; Model 2, CFS which resolved in ten years; Model 3, no CFS but 10% risk of death 10 years later.

**Results:** In all models, acute mortality was only a small share of total morbidity. For lifelong moderate CFS symptoms, healthy years lost per COVID-19 case ranged from 0.92 (male in his 30s) to 5.71 (girl under 10) and were 3.5 and 3.6 for the oldest females and males. At higher symptom severities, young people and females bore larger shares of total morbidity; if symptoms were persistent or survivors’ later mortality increased, young people of both sexes were at highest risk.

**Conclusions:** Compared to post-COVID, acute mortality contributes only a small share of total COVID-19 morbidity. Total burden falls heavily on the young, who are currently deprioritized for preventive interventions such as vaccines. To fairly allocate scarce resources, decisionmakers should consider all morbidity.

## Background

Calculations of disease burden of COVID-19 are used to allocate scarce resources, and generally focus on death and acute illness which are more common in the elderly^1^; thus older patients are prioritized for interventions such as vaccination. Less attention is paid to ‘post-COVID’ or long-term morbidity which follows COVID-19 infection in 10% of cases, of which 80% are female^2^: this translates to 16% of females and 4% of males.

Parallels have been drawn between post-COVID and chronic fatigue syndrome, CFS.^2^ Post-COVID and CFS are both postinfectious syndromes^3^ whose most common symptoms are fatigue, muscle and body aches, and difficulty concentrating;^2,4^ they also both tend to strike women. Significantly, although CFS has been known for decades it remains poorly understood and medically neglected.^2,5^ Diagnosis, treatment and services are not easily accessible even to severe cases; little specific treatment is available; and research is sparsely funded relative to the disease burden.^5^ Much of this also is true for post-COVID, whose prevention should thus be a public health priority.

In addition to CFS-type symptoms, many or most COVID-19 cases have clinical sequelae such as damage to the heart^6^ and lungs,^7^ even in those whose symptoms were otherwise mild.

This damage may increase mortality risk years or decades later, but has not yet been included in calculations of disease burden or even of mortality.

In this paper I establish a plausible range for total morbidity burden per COVID-19 case that is attributable to CFS-type symptoms; to immediate death; and to delayed death. By doing so I inform allocators of scarce resources and suggest avenues for future research.

## Materials and Methods

Disability-adjusted life years (DALY) lost per COVID-19 case were computed for the combination of death and post-COVID by sex and age using sex- and age-specific case fatality rates (CFR)^1^ and estimated remaining lifespan from the US Social Security Administration.^8^ All computations here show DALY lost rather than DALY remaining.

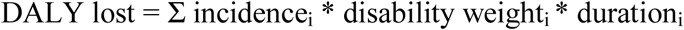

where death has a disability weight of 1.0.

Three models were run:

- Model 1 treated post-COVID as CFS, with increased disability but no increase in mortality. DALY were computed for mild, moderate, and severe CFS (disability weights of 0.14, 0.45, and 0.76)^9^ and presented in a single chart along with those for death.
- Model 2 also treated post-COVID as CFS, but assumed that symptoms resolved ten years after the initial infection.
- Model 3 ignored CFS-type symptoms entirely but assumed that 10% of COVID-19 survivors sustained damage to heart, lungs, or other vital systems which caused death an average of ten years later.

We did not consider post-COVID symptoms which are not also CFS symptoms, such as anosmia, since no published DALY estimates exist for these. However, once these weights become available this issue can be resolved by multiplying together the disability weights:

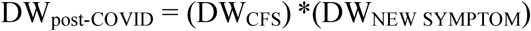

For example, if the disability weight for anosmia was 0.90, and all post-COVID cases had it, the disability weight for mild post-COVID would increase from 0.14 to 0.244.

## Results

These figures show the breakdown of total DALY lost per COVID-19 case under different scenarios for each sex. In models showing symptoms of varying severity, total DALY lost can be read from the top of the colored bar corresponding to the chosen symptom severity.

### Model 1: Persistent Symptoms

If symptoms persisted for life but mortality was not affected (Figure 1), then most morbidity was in female survivors. Female morbidity had a U-shaped association with age, being higher in the young and old (dominated by post-COVID and death, respectively); while male morbidity was J-shaped: dominated by mortality, mostly in the old.

**Figure 1 Caption.**
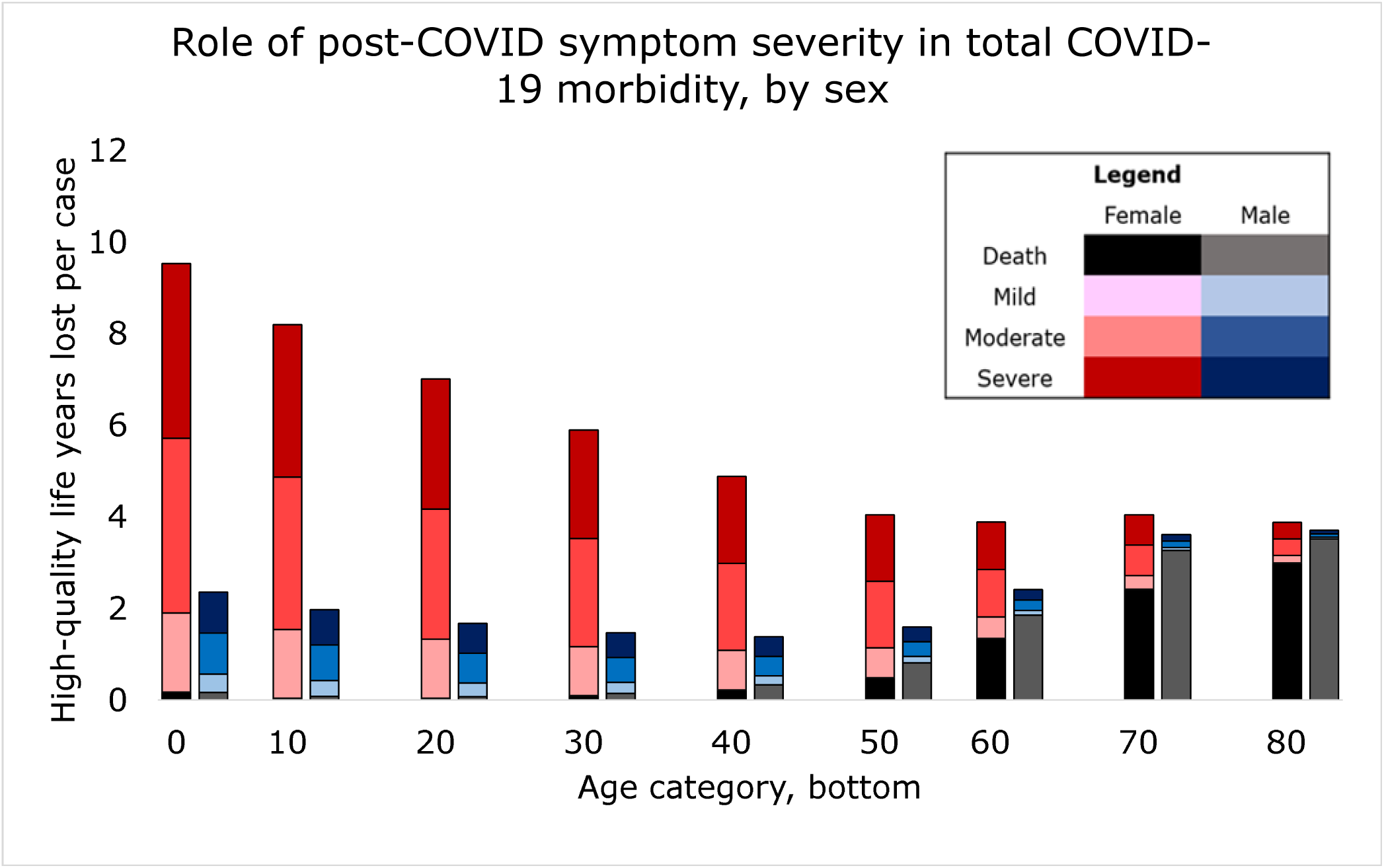
The top of each colored bar segment represents average healthy years lost per COVID-19 case in that group for that severity of post-COVID symptoms (mild, moderate or severe.)

#### Moderate Symptoms

If post-COVID was of moderate severity, each female COVID-19 case under age 10 lost 5.7 DALY, while those over 80 lost 3.5. These can be read from the top of the dark pink bar corresponding to ‘moderate symptoms, female.’ 3% and 86% of these lost DALY were death, while the remainder was disability. Female morbidity was lowest at 2.59 DALY per case (19% death), at age 50-60. Male morbidity in the youngest was 1.5 DALY per case (12% death) it was 0.95 (35% death) at age 40-50, and 3.6 in the oldest (97% death.)

#### Mild Symptoms

Only if symptoms were mild did the oldest females had higher morbidity than the youngest. Girls under 10 lost 1.9 DALY per case (9% death); this declined to 1.08 (21%) in those between 40 and 50, and rose to 3.2 (95%) in the oldest. Male morbidity did not exceed 1 DALY per case until age 60, and then surpassed female to reach a maximum of 3.6. (99%)

#### Severe Symptoms

If post-COVID was severe, females had higher morbidity than males in every age group; and female morbidity dropped near-linearly with age, from 9.5 (2% death) in the youngest to 3.9 (77%) in the oldest. Male morbidity remained dominated by mortality and showed only a slight U-shaped association with age, from 2.4 DALY in the youngest (7% death) to 1-3 in middle age (increasing from 5% death to 77%) and 3.7 (95% death) in the oldest.

### Model 2: Symptoms Resolve

If post-COVID symptoms resolved after ten years (Figure 2), total DALY lost to disability were age-invariant (0.224, 0.720, and 1.216 for mild, moderate and severe symptoms in females, and 0.056, 0.180, and 0.304 for males) until lifespan dropped below ten years. This was age 80 in females and 70 in males. As a result morbidity was almost constant until age 40, when CFR began to increase. Morbidity was about 0.5, 0.8, and 1.3 DALY per case for young females with mild, moderate and severe symptoms (about 40%, 10%, and 5% death) and 0.3, 0.5 and 0.7 for young males (60, 30 and 20% death.)

**Figure 2 Caption.**
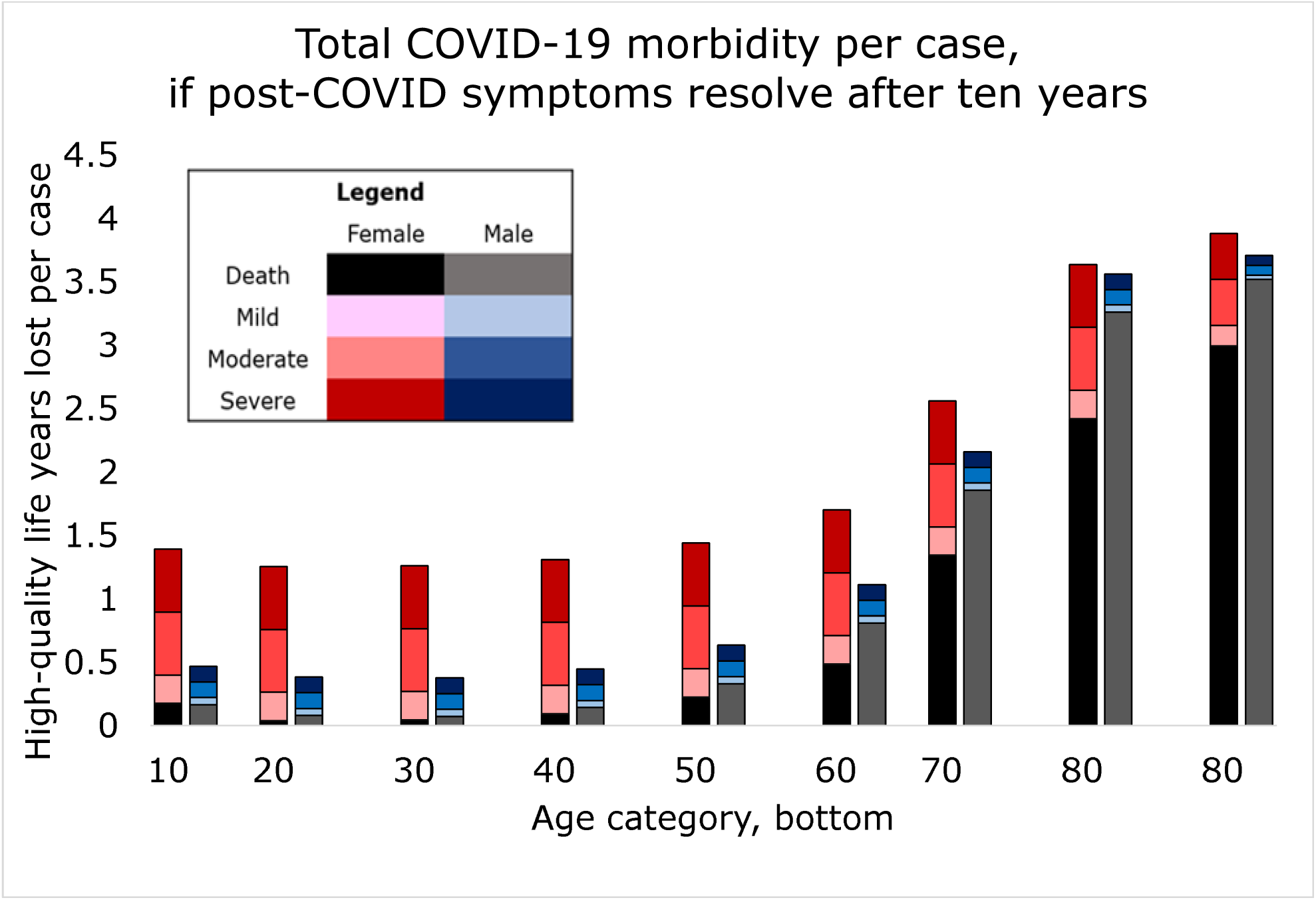
The top of each colored bar segment represents average healthy years lost per COVID-19 case in that group for that severity of post-COVID symptoms (mild, moderate or severe), under the assumption that post-COVID resolves in ten years.

**Figure 3 Caption.**
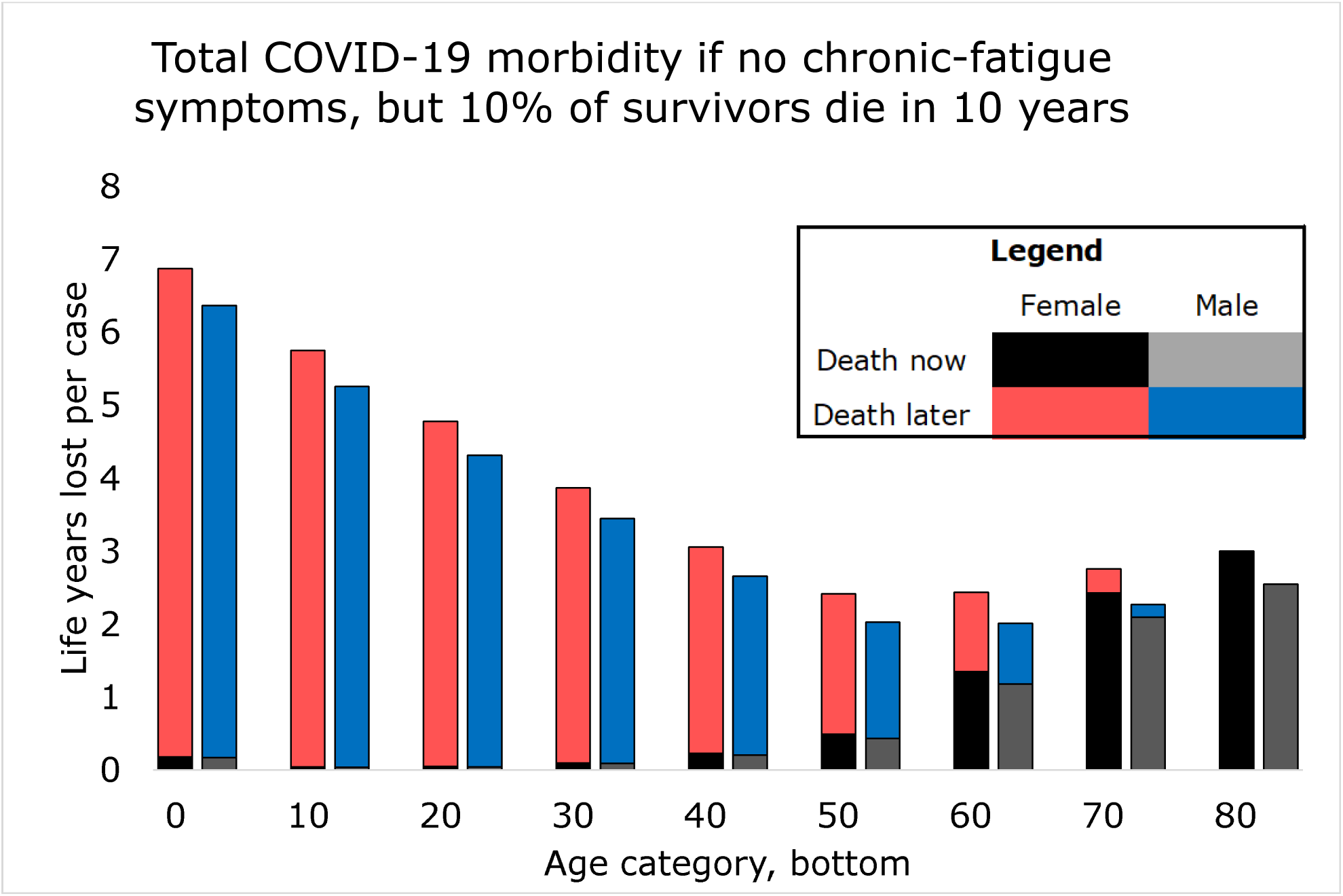
The top of each bar segment represents average life-years lost per COVID-19 case in that group from immediate death (published case fatality) and delayed death an average of 10 years later, such as might be caused by damaged heart or lungs.

### Model 3: Increased Mortality

If 10% of post-COVID cases had symptoms that caused mortality an average of 10 years later, then even if CFS-type symptoms were absent the situation for both sexes was similar to that for females when CFS-type symptoms were severe. Average female cases under 10 in this scenario lost 6.9 DALY, of which 3% were immediate death and 97% were delayed death. Males in this age group lost 6.4 DALY, (3% immediate death). DALY loss for females after age 50 was nearly constant, between 2.4 and 3.1 per case; for males this plateau was earlier (age 40+) and lower (2.0 – 2.7.)

## Discussion

This paper shows that under all but the most optimistic conditions, acute case fatality is likely to contribute only a small share of total COVID-19 morbidity. In most models total burden fell heavily on females and the young. Rather than focusing solely on mortality, allocators of scarce resources should consider all sources of morbidity.

Rather than give a single estimate of the contribution of acute case fatality to total COVID-19 morbidity, this paper provides a plausible range. It is likely that the truth will contain elements of all three models: chronic CFS-type symptoms of varying severity which may resolve, plus elevated mortality in those survivors or others. In determining which scenario is the closest to the truth, researchers should establish post-COVID incidence, both overall and in specific populations; its sex ratio; and its clinical course, which may include remission, death, or some mix.

Estimates of post-COVID incidence vary, but most are higher than the 10% cited by Rubin.^2^ A different paper^10^ found persistent symptoms in a third of outpatients, including some whose initial infection was asymptomatic, up to nine months later; and others found persistent symptoms in half, or more than half, of those who had been hospitalized. ^7,11^ If the true incidence is found to be higher than 10% then the disability burden of post-COVID will increase, equivalent to a worsening of symptoms under the present model; while if persistent symptoms are more common in those with more severe illness, relative burden borne by the old (who tend to have more severe illness) will increase, while that borne by the young (who are more often asymptomatic) will decrease.

Secondly, our models used disability weights for CFS since none exist for post-COVID. However post-COVID has symptoms that CFS does not, leading to systematic underestimation of the true disability weight of post-COVID. For each additional symptom, the number of DALY lost per case at a given symptom severity increases such that the situation becomes similar to that for a higher symptom severity.

Thirdly, the shape of the post-COVID morbidity curve with age is sensitive to the clinical course of the disease. Under most situations the curve was U-shaped (morbidity high in young and old, lower in middle age) or L-shaped (morbidity highest in the young.) These were the situations if post-COVID caused lifelong disability that was other than mild; increased mortality; or both.

However, there was one model in which the mortality curve was J-shaped and burden of morbidity was borne by the elderly (the situation assumed by current public-health guidelines.) For this to occur, non-mild symptoms must be time-limited, resolving either spontaneously or due to medical advances. For either of these to occur, post-COVID would have to be atypical of postinfectious conditions such as CFS. Full recovery from these conditions is rare:^5^ most patients experience fluctuating symptoms, with periods of low and high functioning,^9,12^ and some deteriorate further. Furthermore ‘recovery’ is often defined relative to the disease state rather than relative to fully restored health: even those reported as recovered often have persistent disability.^12^ Thus while post-COVID patients may experience improvements in symptoms, it seems likely that some disability will remain.

It is also possible that medical advances will improve the post-COVID prognosis. However, this would require a significant change in current priorities: relative to its disease burden CFS is deprioritized for research funding^5^ and although it has been documented for almost a century, many patients have difficulty accessing diagnosis, treatment or services. The same is true for post-COVID.^4^ Thus while it is not impossible that post-COVID will become treatable, this scenario appears unlikely.

To conclude, these findings establish plausible outer bounds for the sex and age bias of total disease burden of COVID-19. In most situations, most morbidity is in female survivors and in young people. However, these estimates are imprecise and based on incomplete data. Future research should collect and publish better data to allow fair distribution of resources for prevention of COVID-19 infection; and decisionmakers should allocate those resources to minimize total morbidity, according to the best available knowledge.

## Data Availability

Data are publicly available at the links in the manuscript; worldometers.info

## Acknowledgments and Funding

none

